# Surveillance of Myopericarditis following COVID-19 Booster Dose Vaccination in a Large Integrated Health System

**DOI:** 10.1101/2022.02.08.22270635

**Authors:** Katie A Sharff, David M Dancoes, Jodi L Longueil, Paul F Lewis, Eric S Johnson

## Abstract

**Purpose:** The risk of myopericarditis following COVID-19 booster vaccination has not been extensively evaluated. We provide a timely case ascertainment of myocarditis following COVID-19 booster vaccine in individuals age 18-39 years from an integrated health system.

**Methods:** We studied a cohort of 65,785 Kaiser Permanente (KP) Northwest Health Plan members aged 18-39 years who received a COVID-19 vaccine booster at least 5 months following completion of the primary series. We identified cases of myopericarditis by searching the electronic health record for the National Center for Health Statistics (NCHS) text label for ‘myocarditis’ or ‘pericarditis’ diagnosis codes in all inpatient and outpatient encounters through January 18^th^ 2022. The cohort was followed for 21 days after their booster. We excluded anyone with a documented diagnosis of myocarditis or pericarditis before their first COVID-19 vaccination. Two physicians independently reviewed the identified patient records and applied the CDC myocarditis and pericarditis surveillance case definition to classify records as confirmed, probable or excluded based on the prior published definition.

**Results:** Our method identified 6 patients who met the confirmed or probable CDC case definition for acute myocarditis or pericarditis within 21 days of COVID-19 booster dose among 65,785 eligible members. Four cases occurred in 27,253 men. Overall, we estimated 9.1 cases (exact 95% CI 3.4 to 19.9) of post-booster myopericarditis per 100,000 booster doses given. In men, we estimated 14.7 cases (exact 95% CI 4.0 to 37.6) per 100,000 booster doses given.

**Conclusion:** We identified a rate of 9.1 cases of myopericarditis per 100,000 COVID-19 booster doses which is higher than prior estimates reported by the Vaccine Adverse Event Reporting System (VAERS). Myopericarditis occurs following COVID-19 booster vaccine and may be underreported by current surveillance methods. High sensitivity of these case estimates is essential when modeling risk and benefit for sequential COVID-19 vaccinations for the general population.

## Introduction

COVID-19 vaccine boosters were recommended by the Centers for Disease Control (CDC) for all individuals 18 years and older to provide better protection against circulating variants. The risk of myopericarditis following sequential COVID-19 vaccination needs to be evaluated. We have demonstrated that the Vaccine Safety Datalink (VSD) rapid cycle analysis method identified a lower incidence of myopericarditis following COVID-19 mRNA vaccine, in part because their search for hospital discharge claims omitted ICD-10 codes, and because insurance claims from community hospitals may be delayed by weeks. (1) We provide a more timely and complete case ascertainment of myopericarditis following COVID-19 booster vaccine in individuals aged 18-39 years.

## Methods

We studied a cohort of 65,785 Kaiser Permanente (KP) Northwest Health Plan members aged 18-39 years who received a COVID-19 vaccine booster at least 5 months following completion of the primary series. We identified cases of myopericarditis by searching the electronic health record for the National Center for Health Statistics (NCHS) text label for ‘myocarditis’ or ‘pericarditis’ diagnosis codes in all inpatient and outpatient encounters through January 18^th^ 2022. The cohort was followed for 21 days after their booster. We excluded anyone with a documented diagnosis of myocarditis or pericarditis before their first COVID-19 vaccination. Two physicians independently reviewed the identified patient records and applied the CDC myocarditis and pericarditis surveillance case definition to classify records as confirmed, probable or excluded based on the prior published definition. (2) KP’s institutional review board approved the study.

## Results

Our method identified six patients who met the confirmed or probable CDC case definition for acute myocarditis or pericarditis within 21 days of receipt of COVID-19 booster dose among 65,785 eligible members (Table). Four cases occurred in 27,253 men. All six patients received a Pfizer vaccine as their booster dose. Five out of six patients reported chest pain within 4 days of vaccination, although one patient waited until day 8 to present for her chest pain. Patient #6 developed chest pain, myocarditis, and cardiogenic shock after his booster dose. Though the illness was attributed to the booster dose, his clinicians are concerned that the dose unmasked an underlying autoimmune condition. Additionally, patient #5 presented with mild myocarditis following a heterologous series of Johnson & Johnson vaccine followed by a booster dose of Pfizer vaccine.

**Table:** Summary of Myopericarditis cases Following COVID-19 Booster Dose.

| Case | Age | Sex | Primary Series | Boost Vaccine | Dose | Interval: Prime series to Boost (days) | Chest pain onset (days) | EKG | Trop peak mcg/L | Evaluation of CAD | LVEF on echo, % | LOS, days |
| --- | --- | --- | --- | --- | --- | --- | --- | --- | --- | --- | --- | --- |
| 1 | 18-24 | M | Pfizer | Pfizer | 3 | 204 | 3 | ST elevations | 2.93 | Not done | 55-60% | 2 |
| 2 | 18-24 | M | Pfizer | Pfizer | 3 | 260 | 3 | ST elevation | Trop I 10.4 | Not done | 55% | 1 |
| 3 | 25-29 | M | Pfizer | Pfizer | 3 | 210 | 3 | ST elevations | 10.4 | Not done | 60% | 1 |
| 4 | 30-39 | F | Pfizer | Pfizer | 3 | 239 | 2 (8 days to ED visit) | Diffuse ST changes | < 0.01 | Not done | 55% | 1 |
| 5* | 18-24 | F | J&J | Pfizer | 2 | 196 | 4 | ST abnormality | .04 | Not done | 60-65% | ED |
| 6* | 30-39 | M | Pfizer | Pfizer | 3 | 183 | 19 | Sinus tach | 17.8 | Not done | 35-40% | 4 |

LVEF: Left-ventricular end-systolic function
LOS: Length of Stay
ED: Emergency Department
Normal troponin Range: <=.03 mcg/L
Pfizer: Pfizer BioNTech
J&J: Johnson and Johnson
\*Met CDC case definition, atypical presentation

Overall, we estimated 9.1 cases (exact 95% CI 3.4 to 19.9) of post-booster myopericarditis per 100,000 booster doses given. In men, we estimated 14.7 cases (exact 95% CI 4.0 to 37.6) per 100,000 booster doses given.

## Discussion

We identified a risk of 9.1 cases per 100,000 booster doses. Our small sample size limits the precision of our estimate. This risk is higher than prior estimates reported by Vaccine Adverse Event Reporting System (VAERS) which identified 54 preliminary reports of vaccine related myopericarditis; 12 confirmed and 38 under review, following 26.3 million booster doses administered *across all ages*, with an unadjusted estimate of 0.21 cases per 100,000 doses (95% CI, 0.15 to 0.27). (3) VAERS is passive system relying on patients or providers to report; but limitations include both over and under reporting. (4) Active surveillance by the VSD has not yet reported a risk of myopericarditis following booster vaccinations, although we would anticipate under-estimation due to limitations in their methodology. (1) Israel reported the risk of myopericarditis following booster dose as 4.7 cases per 100,000 in men age 20-24. (5)

Myopericarditis occurs following booster doses and may be underreported by current surveillance methods. Completeness or high sensitivity of these case estimates are essential when modeling risk and benefit for wide-scale vaccine implementation and sequential COVID-19 vaccinations for the general population.

## Data Availability

All data produced in the present study are available upon reasonable request to the authors

